# COVID-19 Impact in Crohn’s Disease Patients Underwent Autologous Hematopoietic Stem Cell Transplantation

**DOI:** 10.1101/2023.06.22.23291610

**Authors:** Milton Artur Ruiz, Roberto Luiz Kaiser Junior, Lilian Piron-Ruiz, Tainara Souza Pinho, Lilian Castiglioni, Luiz Gustavo de Quadros

## Abstract

SARS COV 2 is the virus responsible for COVID-19, a disease that has been blamed for inducing or exacerbating symptoms in patients with autoimmune diseases.

Crohn’s disease (CD) is an inflammatory bowel disease that affects genetically susceptible patients who develop an abnormal mucosal immune response to the intestinal microbiota. Patients who underwent Hematopoietic Stem cell Transplantation are considered at risk for COVID-19.

The objective of this report was to describe for the first time the impact of COVID-19 in a group of 50 patients with Crohn’s Disease (CD, 28 females, and 22 male) with a mean age of 38 years, previously submitted to Autologous, non-myeloablative, Hematopoietic Stem Cell Transplantation (Auto HSCT) between 2013 and 2021. In this series, 19 patients were diagnosed with positive COVID-19. In two (2) patients there was a report of the occurrence of two infectious episodes. Parameters related to HSCT, such as time elapsed since the procedure, vaccination status, CD status before and after infection, and clinical manifestations resulting from COVID-19, were evaluated. Among the patients with COVID-19, in three, submitted to Auto HSCT less than six (6) months ago, there was a change in the CD status, and one of them, in addition to the CD symptoms, started to present thyroid impairment with positive anti-TPO. Only one of the patients required hospitalization for five days to treat COVID-19 and remained in CD clinical remission. Nine patients reported late symptoms that may be related to COVID-19. There were no deaths, and the statistical evaluation of the series of COVID-19 patients after HSCT and those who did not present an infectious episode did not present significant data regarding the analyzed parameters. Despite the change in CD status in three patients and the presence of nine patients with late symptoms, we can conclude that there was no significant adverse impact concerning COVID-19 in the evaluated patients who underwent HSCT to treat CD.

## Introduction

Since 2020, the world has lived with the severe acute respiratory syndrome coronavirus 2 (SARS-CoV-2) pandemic, the disease also known as COVID-19, characterized primarily by respiratory failure and pneumonia but with different repercussions and clinical complications over the entire organism (1,2).

Crohn’s disease (CD) is a chronic disease that occurs in genetically susceptible individuals as a result of an anomalous immune response of the mucosa to the intestinal microbiota (3).

Despite the improvement in the medical management of inflammatory bowel disease (IBD) and CD (CD) in recent years, some patients with CD become refractory or without any therapeutic drug options. Autologous Hematopoietic Stem Cell Transplantation (Auto HSTC) in those patients could be an alternative treatment option (4).

The impact of COVID-19 on Crohn’s Disease and Inflammatory Bowel Diseases is not demonstrated, despite the gut microbiota being considered one of the preferred targets of SARS COV 2 (5).

COVID-19 has been described as triggering or responsible for relapses in patients with autoimmune diseases (6,7).

Hematopoietic stem cell transplant (HSCT) recipients are considered with an increased risk of mortality and morbidity with coronavirus disease 2019 (COVID-19) due to severe immune dysfunction (8).

However, there is no data on the outcomes and the COVID-19 clinical impact of patients with a CD that underwent Auto HSTC.

Based on this data, this report aimed to present and discuss the impact of SARS-COV-2 infection for the first time in patients with severe and refractory CD to conventional treatments who underwent Auto HSCT.

The evaluated patients are part of studies registered in US Clinical Trials NCT 03000296 and IRD-CAAE 20 894719.1.0000.5629, infected or not by SARS-COV 2. All consented to have their data published.

## Patients and Methods

A unicentric longitudinal study was carried out on a group of 50 patients with CD who underwent an Auto HSCT between 2013 and 2021.

Patient information was organized in a database of 50 records and 12 variables. The records contained information on two patients who had the disease twice. The variables obtained from each patient were age, sex, date of the BMT, months elapsed from the transplant until the date of infection by the virus, clinical symptoms observed during the infection, treatment to which the patient Covid 19, history of immunization, the origin of the immunizer, number of doses, patient symptoms during and after COVID 19, in addition to the status of Crohn’s Disease before and after COVID 19.

We consider remission, CD status, and those patients without symptoms or submitted to any time after Auto HSCT for specific clinical or surgical treatment for the disease at the time of this evaluation.

The patient’s time evaluation was in 2022, and the Auto HSCT non-myeloablative regimen data and the details of the procedure they underwent are described in another publication elsewhere (9).

Statistical analysis of patients was organized into two groups, one formed by those who did not have a diagnosis of COVID-19 and the other by those who had a diagnosis of COVID-19. Patients who underwent HSCT after the onset of the pandemic were evaluated separately, as well as those diagnosed with COVID-19, according to the same parameters. Methods of Pearson’s Chi Square test, T test and Fisher exact test were applied in the groups described considering the significance level of 0.05%.

The statistical software used in the analysis was SPSS, version 23. Results

The study consisted of 50 patients of both sexes, 28 (56%) female and 22 (44%) males. The mean age was 38.12 ± 9,273 years, with the youngest patient being 17 and the oldest 57 years old.

The clinical and demographic characteristics of the patients who underwent self-HSCT and contracted COVID-19 are shown in Table 1. In addition, data for the entire series are shown in supplementary data.

Table 2 shows the two groups of patients who had or did not have Covid 19.

Of the total, 31 patients (62%) did not have COVID-19, while 19 (38%) had the diagnosis confirmed by the RT PCR method. Among the patients with COVID-19, 12 (42.8%) were female, seven (31.8%) were male, nine (23.1%) had been previously vaccinated, while 10 (90.9%) still had not taken any doses of the vaccine against Sars-Cov-2.

The immunizer from the Pfizer laboratory predominated about the vaccines applied, with 23 immunized patients, followed by the Astra Zeneca laboratory with 20 patients—45 (90%). Of the patients who reported being vaccinated, 30% received two doses, and 42% received three.

Before the diagnosis of Covid 19, 22 patients were in clinical remission of Crohn’s disease, while 28 were relapsed. After infection, 19 patients were in remission and 31 in relapse. In three patients after Covid 19, there was a change in status, and they started to present CD relapse. After 2019, the beginning of the pandemic, 21 patients underwent Auto HSCT, and the analysis of the same variables as the initial group of 50 patients, mean age was 31 years, a median of 35 years, 11 were female, and 10 were female masculine. In this group, eight had Covid 19 while 13 were negative. Therefore, the analysis results of this group of patients after 2019 are similar and do not differ statistically from the entire group studied.

The symptoms of patients who contracted Covid 19 were those usually described for the disease, such as fever, asthenia, cough, sore throat, and muscle pain, and only one of the patients (#15) required hospitalization for five days.

As additional data, two patients with two infectious episodes (#22 and #40) in the first event were in clinical remission. However, patient # 40, after the first episode, started to show Anti-TPO antibodies in addition to signs of CD activity. The other patient, #22, remained in clinical remission after the two infectious episodes.

Regarding the symptoms the patients had during the infectious episodes, in addition to the usual symptoms, diarrhea was mentioned in 15.8% and vomiting in 15%. After COVID-19, nine patients reported varied late symptoms, and 5.3% reported persistent diarrhea and abdominal pain. Only one of the patients mentioned the presence of cognitive disorders. There were no deaths in that case series related to COVID-19.

The supplementary data in Figure 1 and Tables 3, 4, 5, and 6 complement the data described in the study.

## Discussion

The Sars-Cov-2 virus is responsible for Covid-19, a disease that has impacted the entire world and is blamed for the induction and exacerbation of immunocompromised patients and carriers of autoimmune diseases. (6,7,10).

In addition to immunocompromised patients, HSCT recipients are part of a group of patients at greater risk of contracting COVID-19 (11).

Allogeneic and autologous HSCT recipients have increased mortality, morbidity, and clinical complication rates when contracting COVID-19 (8,12-14). A report on a group of patients undergoing transplantation showed a mortality rate for autologous HSCT of 17%, and for allogeneic HSCT was 21% (8). These data configure the severity to which HSCT recipients are submitted in both modalities. There is a reason for evaluating a group of patients previously submitted to autologous HSCT and its impact on Crohn’s Disease.

Crohn’s Disease courses with dysbiosis of the digestive system, which is the primary habitat and target of infection by Sars-Cov-2. That is due to a high intestinal expression of angiotensin-converting enzyme-2 (ACE2) and serine transmembrane protease-2 (TMPRSS2) receptors, which are found at high levels at this site (15).

The intestinal microbiome of healthy people, compared to that of people with COVID-19 or Crohn’s disease, shows a considerable reduction of anti-inflammatory bacteria (15).

Patients with COVID-19 have gastrointestinal manifestations during the infection, and the Sars-Cov-2 virus is considered to trigger CD in patients predisposed to the disease (16). However, studies on COVID-19 on gut health and inflammation among patients with inflammatory diseases remain uncertain, particularly on the susceptibility and clinical impact of COVID-19 and its impact on the gut microbiota (5).

It is believed that the immune dysregulation caused by COVID-19 can trigger the onset of various autoimmune diseases, including in patients with active CD, and persist after recovery from the infectious episode (17).

COVID-19 shares similarities with autoimmune diseases regarding clinical manifestations, immune responses, and pathogenic mechanisms. In addition, there are reports of several patients with evolution to several autoimmune diseases, such as Guillain Barre Syndrome, lupus, vasculitis, myopathies, and description of isolated cases of multiple sclerosis and Still’s Disease (18-20).

In our series of 50 patients with CD, and in the 19 patients who had COVID-19, there was no description of an increase in the morbidity rate or presence of mortality, data that differ from the literature described, (8, 12-14). In the analysis of the series of patients submitted to autologous and allogeneic HSCT, we observed that they had once-hematological diseases submitted to rescue chemotherapy, mobilization schemes with high doses of chemotherapy, and myeloablative conditioning. Our sample differs in using mobilization regimens with low doses of cyclophosphamide and non-myeloablative conditioning regimen. This may explain the low morbidity, describing only one (1) case that required hospitalization but outside the Intensive Care Unit. Another relevant data observed was the absence of deaths.

In our patients, there was no change in DC status in most cases, with only a change from clinical remission to disease recurrence observed in three cases (#40, #43, and #44). These patients had undergone HSCT less than six months before the infectious episode. This data corroborates the information on patients’ vulnerability in the first year after autologous HSCT (21).

There are reports that COVID-19 can affect the endocrine system and trigger changes in thyroid function, with clinical and biochemical manifestations and the development of antithyroid antibodies. (22,23). One of the cases (#40) that recurred after the infectious episode presented endocrinological manifestations of CD, with the presence of anti-TPO.

Post-COVID syndrome is currently recognized as a new disease in the context of Sars-Cov-2 infection. However, its pathogenesis is still not fully understood, with data that acute inflammation may be responsible for this clinical picture (24-27). Of our patients, nine (9) reported clinical manifestations that may be related to the Post-COVID Syndrome. However, only one (1) of the patients had cognitive disorders, and two (2) patients had digestive manifestations and diarrhea and were in a relapse of the CD.

In conclusion, with the observed data, we can say that the patient with CD who submitted to autologous non-myeloablative HSCT has a low morbidity rate and no mortality and that patients in the first year after transplantation have an increased risk of contracting COVID-19 and with probable repercussions on the status of his illness.

## Data Availability

All data produced in the present work are contained in the manuscript

## Aknowledgements

Camila de Jesus Souza for all support with patients’ data.

