## Supplementary material for "COVID-19 Impact in Crohn’s Disease Patients Underwent Autologous Hematopoietic Stem Cell Transplantation": Figure 1

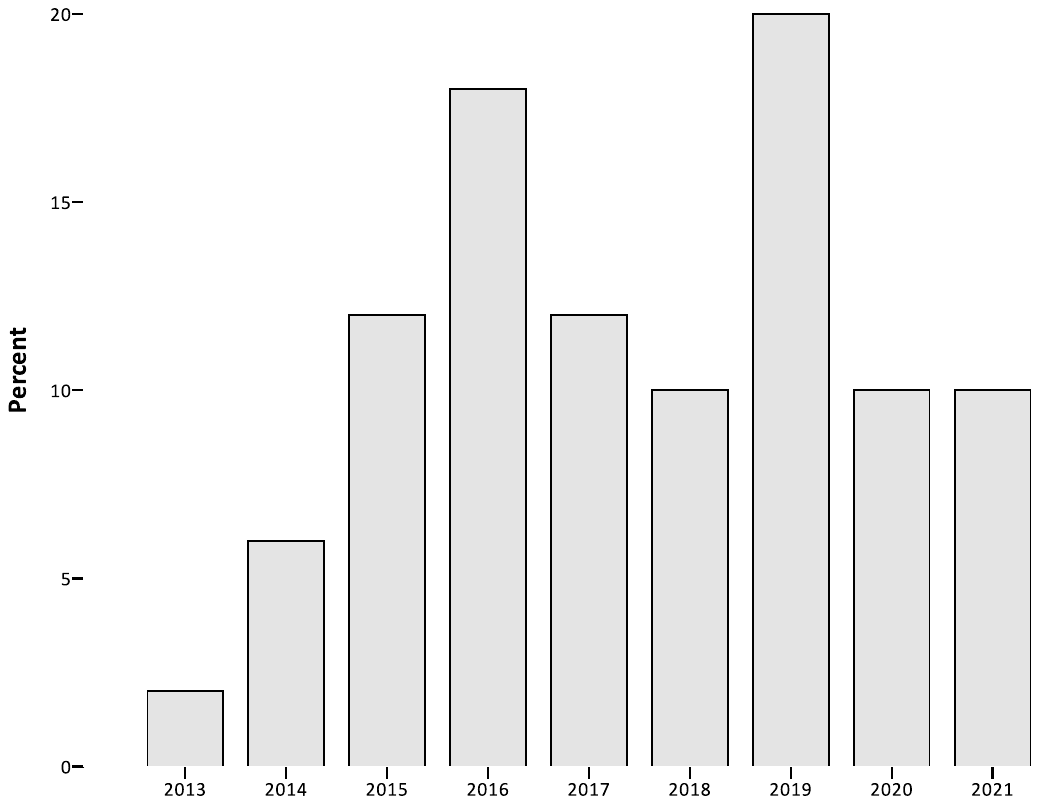

**Year of TMO**

**Figure 1 – Graphic Annual Distribution of Autologous Hematopoietic Stem Cell Transplantation (50 pts – 2013-2021)**
