## Supplementary material for "COVID-19 Impact in Crohn’s Disease Patients Underwent Autologous Hematopoietic Stem Cell Transplantation": Table 2

| **Table 2 - General Statistics of the Crohn Disease Patients Underwent to Autologous Hematopoietic Stem Cell Transplantation and COVID 19 (50 pts. 2013-2021)** | | | | | | | |
| --- | --- | --- | --- | --- | --- | --- | --- |
|  | | COVID 19 Positive Patients  19 (38 %) | | COVID 19 Negative Patients  31 (62 %) | | |  |
| Sex | Female | 12(42.85 %) | | 16(57.14%) | | | 0.425 ^a^ |
|  | Male | 7(31.81 %) | | 15(68.18 %) | | |  |
| Age |  | 39.11±10.104 | | 37.52±8.84 | | | 0.562 ^b^ |
| Elapsed Time HSCT until Evaluation (Months) |  | 46.05±24.19 | | 48.71±28.12 | | | 0.734 ^b^ |
| Elapsed Time HSCT until Covid 19 + |  | 39.47±26.57 | |  | | |  |
| COVID 19  Vaccination | Yes | 9 (23.1 %) | | 30 (76.9 %) | | |  |
|  | No | 10 (90.90 %) | | 1 (9.1 %) | | |  |
| CD status before  COVID 19 | Relapsed | 8 (28.57 %) | | 20(71.42 %) | | | 0.1500^c^ |
|  | Remission | 11 (50 %) | | 11 (50 %) | | |  |
| CD status after  COVID 19 | Relapsed | 11 (35.48 %) | | 20 (64.52 %) | | | 0.7661^c^ |
|  | Remission | 8 (42.10 %) | | 11 (57.89 %) | | |  |

^a^ Test Qui Square Pearson, ^b^ T Test, ^c^ Fisher's Exact Test
