## Supplementary material for "COVID-19 Impact in Crohn’s Disease Patients Underwent Autologous Hematopoietic Stem Cell Transplantation": Table 3

| **Table 3 - Demographic and Clinical Characteristics of Crohn Disease Patients Underwent Autologous HSCT ( 2013 -2021 - 50 pts)** | | | | | | | | | | | | | | | |
| --- | --- | --- | --- | --- | --- | --- | --- | --- | --- | --- | --- | --- | --- | --- | --- |
| **#** | **Age** | **Sex** | **HSCT Date** | **Elapsed Months** | **Vaccina** | **( n ) label vaccine** | **CD Status** | **COVID 19** | **Months HSCT to COVID** | **Vaccination at infection time** | **Symptoms COVID 19 +** | **Treatment COVID 19+** | **CD Status before COVID 19 pts +** | **CD Status after COVID-19 pts +** | **Delayed Symptoms after COVID 19** |
| **1** | 31-35 | F | 2013 | 100 | yes | (n) | Relapsed | no |  |  |  |  |  |  |  |
| **2** | 56-60 | F | 2014 | 88 | yes | (2) Az | Relapsed | no |  |  |  |  |  |  |  |
| **3** | 31-35 | M | 2014 | 88 | yes | (2) Az (1) Pf | Remission | no |  |  |  |  |  |  |  |
| **4** | 41-45 | F | 2014 | 86 | yes | (3) Pf | Relapsed | no |  |  |  |  |  |  |  |
| **5** | 41-45 | F | 2015 | 84 | no | 0 | Remission | yes | 76 | no | Fever, flu-like symptoms, cough, body aches | 0 | Remission | Remission | 0 |
| **6** | 31-35 | F | 2015 | 82 | yes | (2) Pf | Recidiva | no |  |  |  |  |  |  |  |
| **7** | 56-60 | M | 2015 | 78 | yes | (2) Az | Remission | yes | 77 | yes | Fever, body aches, fatigue | 0 | Remission | Remission | 0 |
| **8** | 51-55 | M | 2015 | 77 | no | 0 | Remission | yes | 76 | no | Severe body aches, severe headache, physical and mental fatigue and diarrhea. | 0 | Remission | Remission | Fatigue, muscle weakness, shortness of breath |
| **9** | 46-50 | M | 2015 | 77 | yes | (1) Ja | Relapsed | no |  |  |  |  |  |  |  |
| **10** | 46-50 | F | 2015 | 76 | yes | (2) Az (1) Pf | Remission | no |  |  |  |  |  |  |  |
| **11** | 26-30 | M | 2016 | 73 | yes | (2) Az (1) Pf | Relapsed | no |  |  |  |  |  |  |  |
| **12** | 31-35 | M | 2016 | 71 | no | 0 | Relapsed | yes | 59 | no | flu symptoms | 0 | Relapsed | Relapsed | 0 |
| **13** | 36-40 | M | 2016 | 69 | yes | (2) Ja | Remission | no |  |  |  |  |  |  |  |
| **14** | 41-45 | M | 2016 | 69 | yes |  | Relapsed | yes | 68 | yes | Coryza and sore throat | 0 | Relapsed | Relapsed | 0 |
| **15** | 51-55 | F | 2016 | 69 | yes | 0 | Remission | yes | 68 | no | Headache, diarrhea Hospital stay 5 days | Iver, Cort e Azitro | Remission | Remission | Sleep disorders, headache |
| **16** | 36-40 | F | 2016 | 68 | yes | (2) Az (1) Pf | Remission | no |  |  |  |  |  |  |  |
| **17** | 26-30 | F | 2016 | 67 | yes | (2) Az | Relapsed | no |  |  |  |  |  |  |  |
| **18** | 31-35 | F | 2016 | 65 | yes | (2) Az (1) Pf | Relapsed | no |  |  |  |  |  |  |  |
| **19** | 26-30 | M | 2017 | 59 | yes | (2) Az | Relapsed | no |  |  |  |  |  |  |  |
| **20** | 31-35 | F | 2017 | 57 | yes | (2) Az (1) Pf | Remission | yes | 56 | yes | Headache, rashes, fever, cough and sore throat | Azitro, Cort | Remission | Remission | Loss of taste, nausea and alopecia |
| **21** | 41-45 | F | 2017 | 56 | yes | (2) Az (1) Pf | Remission | no |  |  |  |  |  |  |  |
| **22a** | 26-30 | F | 2017 | 55 | no | 0 | Remission | yes | 41 | no | Flu-like symptoms nausea, fatigue | 0 | Remission | Remission | 0 |
| **22b** | 26-30 | F | 2017 | 55 | yes | (2) Az | Remission | yes | 54 | yes | cough | 0 | Remission | Remission | Diarrhea |
| **23** | 36-40 | F | 2017 | 51 | yes | (2) Az (1) Pf | Relapsed | no |  |  |  |  |  |  | Abdominal pain, Constipation, Bleeding stool |
| **24** | 41-45 | F | 2017 | 50 | yes | (2) Az (1) Pf | Relapsed | yes | 49 | yes | Fever, sore throat, body aches, vomiting, headache, cough and worsening diarrhea. | 0 | Relapsed | Relapsed | Cough, headache and dizziness, burning in the throat |
| **25** | 41-45 | M | 2018 | 48 | yes | (2) Az (1) Pf | Remission | no |  |  |  |  |  |  |  |
| **26** | 51-55 | F | 2018 | 47 | yes | (2) Az (1) Pz | Remission | no |  |  |  |  |  |  |  |
| **27** | 41-45 | F | 2018 | 45 | yes | (1) Az | Remission | yes | 32 | no | Body ache and sore throat. | Azitro, Iver | Remission | Remission | 0 |
| **28** | 31-35 | M | 2018 | 43 | yes | (4) Pf | Relapsed | no |  |  |  |  |  |  |  |
| **29** | 36-40 | F | 2018 | 42 | yes | (2) Az (1) Pf | Relapsed | yes | 41 | yes | Sore throat, cough, runny nose, body ache, fever, indisposition. | 0 | Relapsed | Relapsed | 0 |
| **30** | 51-55 | F | 2019 | 35 | yes | (3) Pf | Relapsed | no |  |  |  |  |  |  |  |
| **31** | 36-40 | M | 2019 | 33 | yes | 0 | Relapsed | yes | 24 | yes | Shortness of breath, cough and asthma | Azitro, Cort | Relapsed | Relapsed | Coagnitive disorders |
| **32** | 26-30 | M | 2019 | 32 | yes | (2) Co | Remission | no |  |  |  |  |  |  |  |
| **33** | 16-20 | F | 2019 | 32 | no | 0 | Relapsed | yes | 12 | no | Headache, sneezing, body ache, runny nose | 0 | Relapsed | Relapsed | 0 |
| **34** | 26-30 | F | 2019 | 30 | yes | (3) Pf | Relapsed | yes | 29 | yes |  | 0 | Relapsed | Relapsed | 0 |
| **35** | 41-45 | M | 2019 | 29 | yes | (2) Az | Relapsed | no |  |  |  |  |  |  |  |
| **36** | 41-45 | F | 2019 | 29 | yes | (2) Az (1) Pf | Remission | no |  |  |  |  |  |  |  |
| **37** | 46-50 | F | 2019 | 28 | yes | (2) Az (1) Co | Relapsed | no |  |  |  |  |  |  |  |
| **38** | 21-25 | M | 2019 | 27 | yes | (3) Pf | Relapsed | no |  |  |  |  |  |  |  |
| **39** | 31-35 | F | 2019 | 26 | yes | (2) Az (1) Co | Relapsed | no |  |  |  |  |  |  |  |
| **40a** | 26-30 | F | 2020 | 24 | no | 0 | Relapsed | yes | 6 | no | Loss of taste and smell, Fever, tiredness, diarrhea, sore throat, headache and body pain.Loss of taste and smell, fever, tiredness, diarrhea, sore throat, headache and body pain. | 0 | Remission | Relapsed | Muscle weakness, sleep and cognitive disturbances with CD symptoms. |
| **40b** | 26-30 | F | 2020 | 24 | yes | (2) Az (1) Pf | Relapsed | yes | 23 | yes | sore throat, cough, runny nose, fever, headache and body ache | 0 | Relapsed | Relapsed | Muscle weakness, sleep and cognitive disturbances with CD symptoms. |
| **41** | 51-55 | F | 2020 | 24 | yes | (2) Pf | Relapsed | yes | 23 | yes | Fever, body aches, sore throat and weakness | 0 | Relapsed | Relapsed | Diarrhoea, severe abdominal pain |
| **42** | 31-35 | M | 2020 | 23 | yes | (2) Az | Relapsed |  | no |  |  |  |  |  |  |
| **43** | 46-50 | M | 2020 | 16 | yes | 0 | Relapsed | yes | 6 | no | Fever, stuffy nose, loss of smell and taste, and muscle aches | 0 | Remission | Relapsed | 0 |
| **44** | 36-40 | M | 2020 | 14 | yes | 0 | Relapsed | yes | 4 | no | Fever, headache, lack of appetite, tiredness, cough, weakness. | 0 | Remission | Relapsed | 0 |
| **45** | 26-30 | F | 2021 | 11 | yes | (2) Az (1) Pf | Remission | no |  |  |  |  |  |  |  |
| **46** | 46-50 | M | 2021 | 8 | yes | (2) Pf | Relapsed | no |  |  |  |  |  |  |  |
| **47** | 31-35 | M | 2021 | 8 | yes | (2) Pf | Relapsed | no |  |  |  |  |  |  |  |
| **48** | 36-40 | M | 2021 | 6 | no | 0 | Relapsed | no |  |  |  |  |  |  |  |
| **49** | 26-30 | M | 2021 | 5 | yes | (2) Az | Remission | no |  |  |  |  |  |  |  |
| **50** | 31-35 | F | 2021 | 5 | yes | (2) Az (1) Pf | Remission | yes | 3 | yes | Fever, headache, body ache, sore throat, cough, tiredness, chest pain, vomiting, diarrhea. | 0 | Remission | Remission | 0 |
| Az: Oxford/Astra Zeneca ChAdOx1-S, Azitro: Azytromicin, Co: Coronavac ChAdOx1 nCOV-19; Cort: Corticosteroids F: female; Iver: Ivermectin; Ja: Janssen SARS COV 2 Ad26-COV2- S; M: male; Pf: Pfizer Biontech BNT16-2b2. | | | | | | | | | | | | | | | |
| Line gris: Covid 19 positive patients | | | | | | | | | | | | | | | |
