## Supplementary material for "COVID-19 Impact in Crohn’s Disease Patients Underwent Autologous Hematopoietic Stem Cell Transplantation": Table 4

**Table 4 – Frequency annual distribution, Autologous Hematopoietic Stem Cell Transplantation** **(50 pts 2013- 2021)**

|  | | Frequency | Percent |
| --- | --- | --- | --- |
|  | 2013 | 1 | 2.0 |
|  | 2014 | 3 | 6.0 |
|  | 2015 | 6 | 12.0 |
|  | 2016 | 9 | 18.0 |
|  | 2017 | 6 | 12.0 |
|  | 2018 | 5 | 10.0 |
|  | 2019 | 10 | 20.0 |
|  | 2020 | 5 | 10.0 |
|  | 2021 | 5 | 10.0 |
