## Supplementary material for "COVID-19 Impact in Crohn’s Disease Patients Underwent Autologous Hematopoietic Stem Cell Transplantation": Table 5

**Table 5. Descriptive Statistics of the Time Elapsed Between the Autologous Hematopoietic Stem Cell Transplantation and Evaluation Date (50 pts. 2013 - 2021)**

|  | | Elapsed Months until Auto HSCT |
| --- | --- | --- |
| Mean |  | 47.70 |
| Median | | 47.50 |
| Mode | | 69 |
| Std. Deviation | | 26.478 |
| Minimum | | 5 |
| Maximum | | 100 |
| Percentiles | 25 | 26.75 |
|  | 75 | 69.50 |
