## Supplementary material for "COVID-19 Impact in Crohn’s Disease Patients Underwent Autologous Hematopoietic Stem Cell Transplantation": Table 6

**Table 6. Descriptive statistics of the Time Elapsed Between the Autologous Hematopoietic Stem Cell Transplantation of the COVID + Patients and Infection Date.**

|  | | Elapsed Months until HSCT to COVID 19 + |
| --- | --- | --- |
| Mean |  | 39.47 |
| Mean | | 39.47 |
| Median | | 41.00 |
| Mode | | 6^a^ |
| Std. Deviation | | 26.578 |
| Minimum | | 3 |
| Maximum | | 77 |
| Percentiles | 25 | 12.00 |
|  | 75 | 68.00 |
